# Pharmacogenomics of GLP-1 Receptor Agonists: A genome wide analysis of observational data and large randomized controlled trials

**DOI:** 10.1101/2022.05.27.22271124

**Authors:** Adem Y Dawed, Andrea Mari, Timothy J McDonald, Lin Li, Shuaicheng Wang, Mun-Gwan Hong, Sapna Sharma, Neil R Robertson, Anubha Mahajan, Xuan Wang, Mark Walker, Stephen Gough, Leen M ‘t Hart, Kaixin Zhou, Ian Forgie, Hartmut Ruetten, Imre Pavo, Pallav Bhatnagar, Angus G Jones, Ewan R Pearson, DIRECT consortium

## Abstract

**Background:** In the treatment of type 2 diabetes (T2D), GLP-1 Receptor Agonists (GLP-1RA) lower glucose levels and body weight, and have cardiovascular benefits. GLP-1RA efficacy and side effects vary between people. Human pharmacogenomic studies of this inter-individual variation can provide both biological insight into drug action and provide biomarkers to inform clinical decision making. We therefore aimed to identify genetic variants associated with glycaemic response to GLP-1RA treatment.

**Methods:** We studied HbA1c reduction at 6 months after starting GLP-1RA in 4,571 subjects with T2D from four prospective observational cohorts and two randomized clinical trials. We evaluated variants in *GLP-1R*, then undertook a genome-wide association study (GWAS) and gene-based burden test.

**Findings:** Variation in HbA1c reduction with GLP-1RA treatment was associated with rs6923761G>A (Gly168Ser) in the *GLP-1R* (0.9 mmol/mol lower reduction in HbA1c per Serine, p=6.0×10^−05^) and low frequency variants in *ARRB1* (pskato=6.72×10^−08^), largely driven by rs140226575G>A (Thr370Met) (2.7mmol/mol greater HbA1c reduction per Methionine, p=5.2×10^−06^). A similar effect size for the *ARRB1* Thr370Met was seen in Hispanic and American Indian populations who have a higher frequency of this variant (6-11%) than in White populations. A genetic risk score derived from these two genes identified around 5% of the population who had a ∼30% greater reduction in HbA1c than the ∼43% of the population with the worse response.

**Interpretation:** This first genome wide pharmacogenomic study of GLP-1RA has provided novel biological and clinical insights. Clinically, when genotype is routinely available at the point of prescribing, individuals with *ARRB1* variants may benefit from earlier initiation of GLP-1RA.

**Funding:** Innovative Medicines Initiative, Wellcome Trust

## Introduction

In the ADA/EASD guidelines, GLP-1 Receptor Agonists (GLP-1RA) are recommended as a second-line and third-line therapy option in people with T2D across the treatment continuum, including those at high risk of or with established atherosclerotic cardiovascular disease, chronic kidney disease, or heart failure and in patients without these comorbidities who have a compelling need to reduce the risk of hypoglycemia, minimize weight gain, or promote weight loss.^1^ There is considerable heterogeneity in how well patients respond to GLP-1RA treatment, with some individuals achieving marked response and others achieving no improvement.^2,3^ However, the mechanisms for this variation are uncertain.

Glycaemic response to metformin and sulphonylureas treatment has been established to be heritable, with 34%-37% of the variance explained by common genetic variants.^4,5^ Pharmacogenomic studies have the potential to reveal novel insights into drug action, reflecting variation in underlying pharmacokinetics (e.g. *SLCO1B1/3* variation for Sulphonylureas)^4^, or pharmacodynamic variation due to variation specific to the drug target and downstream mechanisms, or more generally due to variation in diabetes aetiology (e.g. *ATM/NPAT* or *SLC2A2* variation for metformin).^5,6^ We hypothesize that pharmacogenomic studies of GLP-1RA would provide similar insights into their mechanism of action and have the potential to identify variants that could be clinically informative for treatment decisions. We therefore studied the impact of common, low frequency and rare genetic variants on glycaemic response to GLP-1RA in 4,571 patients with T2D as part of the IMI funded DIabetes REsearCh on patient straTification (DIRECT) Consortium, including genome wide data from two large randomised controlled trials.

## Methods

### Study design and participants

DIRECT is a pan-European consortium established with the overarching aim to identify and validate biomarkers that address current bottlenecks in diabetes drug development and to develop a stratified medicines approach to the treatment of T2D with either existing or novel therapies.^7,8^ In these analyses, we included 4 prospective observational studies consisting of 1,238 participants with T2D of European ancestry (DIRECT, PRIBA (Predicting Response to Incretin Based Agents in Type 2 Diabetes), PROMASTER (PROspective Cohort MRC ABPI STratification and Extreme Response Mechanism in Diabetes) and GoDARTS (Genetics of Diabetes Audit and Research in Tayside Scotland)), and two clinical trial datasets from the HARMONY trials (RCTs of Albiglutide, Supplementary Tabe 1) and AWARD trials (RCTs of Dulaglutide, Supplementary Table 2) consisting of 3,333 participants randomized to the GLP-1RA group. Finally, the GLP-1RA arm of the HARMONY outcomes data consisting of 3,748 participants was included for top hit replication and ethnicity stratified analysis. Detailed information on each cohort and the inclusion criteria is provided in the Supplementary document. All human research was approved by the relevant institutional review boards, and all participants provided written informed consent.

### Genotyping and imputation

A full description of each dataset and the quality control (QC) procedures are available in the Supplementary document. In short, each dataset underwent initial QC and imputation. Standard QC procedures were applied to all the data sets to filter poorly performing genetic markers and samples prior to imputation. Each study performed imputation to the 1000 Genomes reference panel. Post-imputation, variants with poor imputation quality (Info < 0.3 for common variants and <0.6 for low frequency and rare variants) and monomorphic variants were excluded.

### Assessment of glycaemic response to GLP-1RA

Glycaemic response to GLP-1RA was assessed as the quantitative phenotype of HbA1c reduction between baseline HbA1c and treatment HbA1c while the patients were maintained on stable treatment. The baseline HbA1c was the closest HbA1c measure within 3 months prior to GLP-1RA start whilst treatment HbA1c was taken as the closest HbA1c measure to 6 months and within 3-9 months after treatment initiation.

### Statistical analyses

We first performed a candidate gene analysis for two variants in the *GLP-1R* (rs6923761(Gly168Ser), rs10305420 (Pro7Leu)): then two genome wide studies, a GWAS on common variants (MAF ≥5%) and gene-based burden tests on low frequency and rare coding variants (MAF < 5%). For all analyses, we used a linear regression model with HbA1c reduction as the dependent variable, adjusted for baseline HbA1c, sex, duration of diabetes, principal components and other study specific covariates. Baseline HbA1c has been shown as the strongest predictor of glycaemic response in pharmacogenomic studies in diabetes^9,10^ and our aim was to discovery variants that altered treatment HBA1c independent of baseline HbA1c and therefore baseline HBA1c was included as a covariate in all the cohorts.

### Candidate gene analysis

Given the role of the GLP-1R as the target for GLP-1RA, we examined two non-synonymous variants in the *GLP-1R* gene, rs6923761G>A(Gly168Ser) and rs10305420C>T (Pro7Leu), which have previously been reported to potentially alter response to GLP-1 in hyperglycaemic clamp studies^11-16^. The association between these variants and glycaemic response to GLP-1RA treatment merits study in large populations. In this study, we assumed an additive genetic model. Study specific estimates were combined with fixed-effect meta-analysis using the metaphor package in R.^17^ Accounting for the two tests, p ≤ 0.025 was considered as statistically significant.

### Genome-wide association analysis

For the meta-GWAS, each respective cohort conducted GWAS under an additive genetic model to assess the role of common variants (MAF ≥ 5%) in glycemic response to GLP-1RA. Each variant was tested for association with GLP-1RA related HbA1c reduction within each cohort. A linear regression model was undertaken as outlined above, including principal components as covariates; post-regression QC of the summary statistics was performed using EasyQC R package^18^. Post-regression, variants with an imputation quality score <0.3 or absolute beta or standard error >10 were removed. All datasets were meta-analyzed using an inverse variance weighted fixed effect model, implemented in GWAMA (https://github.com/Kyoko-wtnb/mvGWAMA). Post meta-analysis, variants available in less than four studies were excluded. We used the commonly accepted threshold of 5.0×10^−08^ for joint p values to determine statistical significance. Cochran’s heterogeneity statistic’s p-value was reported as Phet. The CMplot package^19^ in R was used to generate Manhattan and quantile-quantile plots.

### Rare variant analysis

To understand the impact of less common and rare coding variants on glycaemic response to GLP-1RA, we performed gene-based burden and variance-component association tests (SKAT/SKAT-O) that were designed to improve power for a mixed set of risk and protective variants on lower frequency and rare protein coding variants (MAF < 5%) across the DIRECT, PRIBA, HARMONY and AWARD cohorts. The R package seqMeta (v1.6.7) was used for these analyses (https://github.com/DavisBrian/seqMeta). seqMeta Computes necessary information to meta analyze gene-based tests for genetic variants in individual studies and performs meta-analysis. We used the burden and dispersion-based association methods. The burden test collapsed genetic variants for each gene by means of a count statistic, which is defined by the number of rare alleles in the gene region. Weights for SKAT were chosen as proposed by Wu et al^20^. Burden and SKAT tests were performed using a quantitative measure of HbA1c reduction in the linear regression model outlined above. To correct for multiple testing, a p value less than 3.66×10^−06^ (corresponding to 0.05/13,654 tested genes) was considered statistically significant. Top hit replication in the *ARRB1* gene was performed using data from the HARMONY outcomes trial.

### Sensitivity analyses

Genetic variants that are significantly associated with HbA1c reduction in the above analyses were also tested for their association with weight reduction after six months of treatment with GLP-1RA using data from the HARMONY phase 3 trials, DIRECT, PRIBA and PROMASTER (n=3009). Association between identified variants and treatment response to metformin and sulphonylureas has also been investigated using our previously published data from the MetGen and MetGen Plus consortiums.^4,6^ In addition, we investigated association between identified variants and HbA1c reduction using data from other comparator arms in the HARMONY trials. Finally, we generated a genetic risk score (GRS) based on the *GLP-1R* and *ARRB1* variants and the GRS was tested against HbA1c reduction after adjusting for the same covariates as in the GWAS.

## Results

### Participant characteristics

We included 4571 individuals with T2D with complete data for HbA1c reduction at 6 months and the covariates. The baseline data for the four observational studies and two RCTs are provided in Supplementary Table 3. Women constitute around 45% of the participants, the mean age range across studies was 54-59 years. Baseline HbA1c varied from 8.14% (65.5 mmol/mol) in the AWARD studies to 9.85% (84.2 mmol/mol) in PROMASTER. The mean HbA1c reduction at 6 months ranges from 0.75% (8.11 mmol/mol) in GoDARTS to 1.38% (15.1 mmol/mol) in PRIBA.

### The serine variant at Gly168Ser in the GLP-1R reduces glycaemic response to GLP-1RA

We investigated the association between two common missense variants, rs6923761G>A (Gly168Ser, MAF = 0.27) and rs10305420C>T (Pro7Leu, MAF = 0.32), and HbA1c reduction after GLP-1RA treatment. In a meta-analysis consisting of 4,571 individuals, Gly168Ser was significantly associated with glycemic response to GLP-1RA. Each copy of the Ser-allele was associated with a lower HbA1c reduction of -0.08%(0.9mmol/mol) (p=6.0×10^−05^, phet=0.20). This association was consistent across the constituent datasets of the meta-analysis (Figure 1). There was no association between Pro7Leu and HbA1c reduction (ß =-0.04% (0.4 mmol/mol)) (p =0.11, phet=0.13).

**Figure 1:**
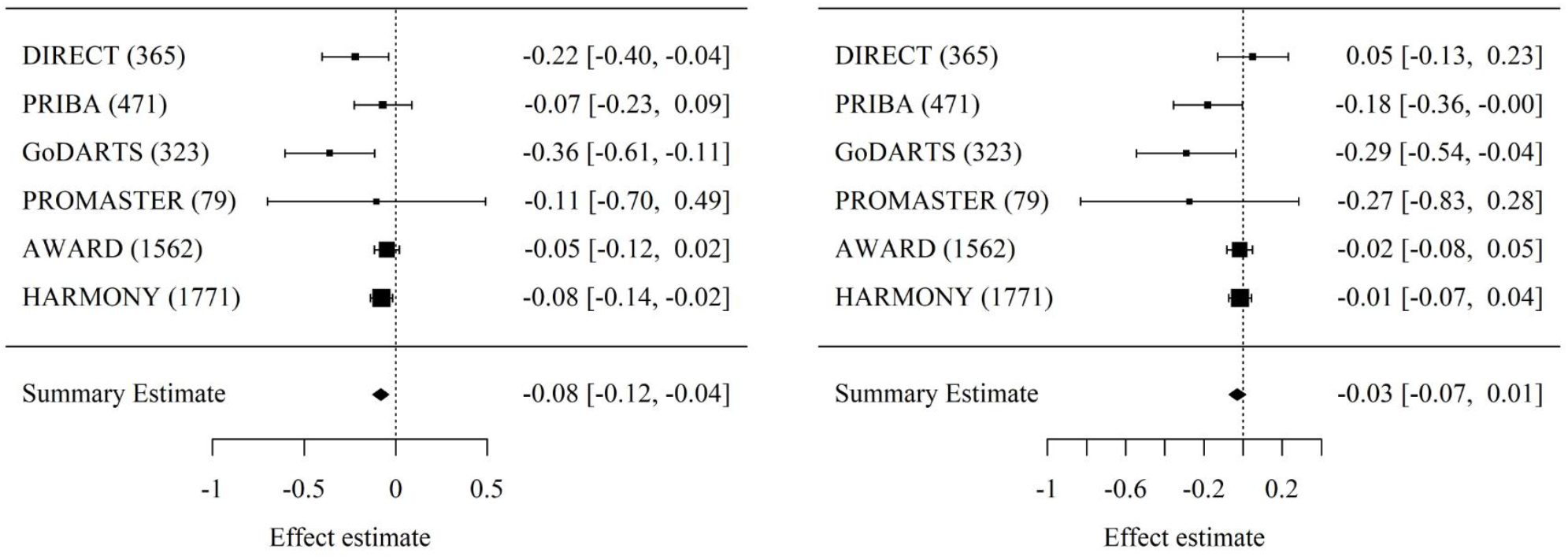
Forest plot representing meta-analysis of the association between *GLP1R*, Gly168Ser (left) and Pro7Leu (right) and HbA1c reduction observed across DIRECT, PRIBA, GoDARTS, PROMASTER, AWARD and HARMONY studies. Effect estimates represent HbA1c reduction (DCCT) per minor allele. Study sample size is shown in parentheses.

### Genome wide association study on common variants revealed no genome-wide significant association

The meta-GWAS across four prospective observational cohorts and two trials included 6,228,682 common autosomal variants from 4,571 independent individuals treated with GLP-1RA (Supplementary Figure 1). No inflation of variant test statistics was observed on quantile-quantile plots and estimates of the genomic inflation factor (λ = 1.04 (Supplementary Figure 2). There was no association for any common genetic variant with HbA1c reduction with GLP-1RA treatment at the genome-wide level of statistical significance (p<5×10^−08^). However, multiple implicated loci with lead variant/nearest gene at rs61800555/*LMX1A*, rs2268640/*GLP-1R*, rs7687008/*LINC00499* and rs10224036/*PTPN12*, reached a suggestive genome-wide significance level (p<1×10^−05^; Supplementary Figure 1, Supplementary Table 4).

### Low frequency and rare variants in ARRB1 are associated with glycaemic response to GLP-1RA

In the gene based meta-analysis there was one gene, Arrestin beta 1 (*ARRB1*), for which the burden of low frequency and rare coding genetic variants was associated with HbA1c reduction (cumulative MAF of 2.4%, pburden=1.12×10^−07^, pskat-o=6.72×10^−08^) at the genome-wide level of statistical significance [inverse variance weighted (IVW) meta-analysis P < 3.66×10^−06^, a Bonferroni correction for 13,654 genes] (Supplementary Table 5, Figure 2).

**Figure 2:**
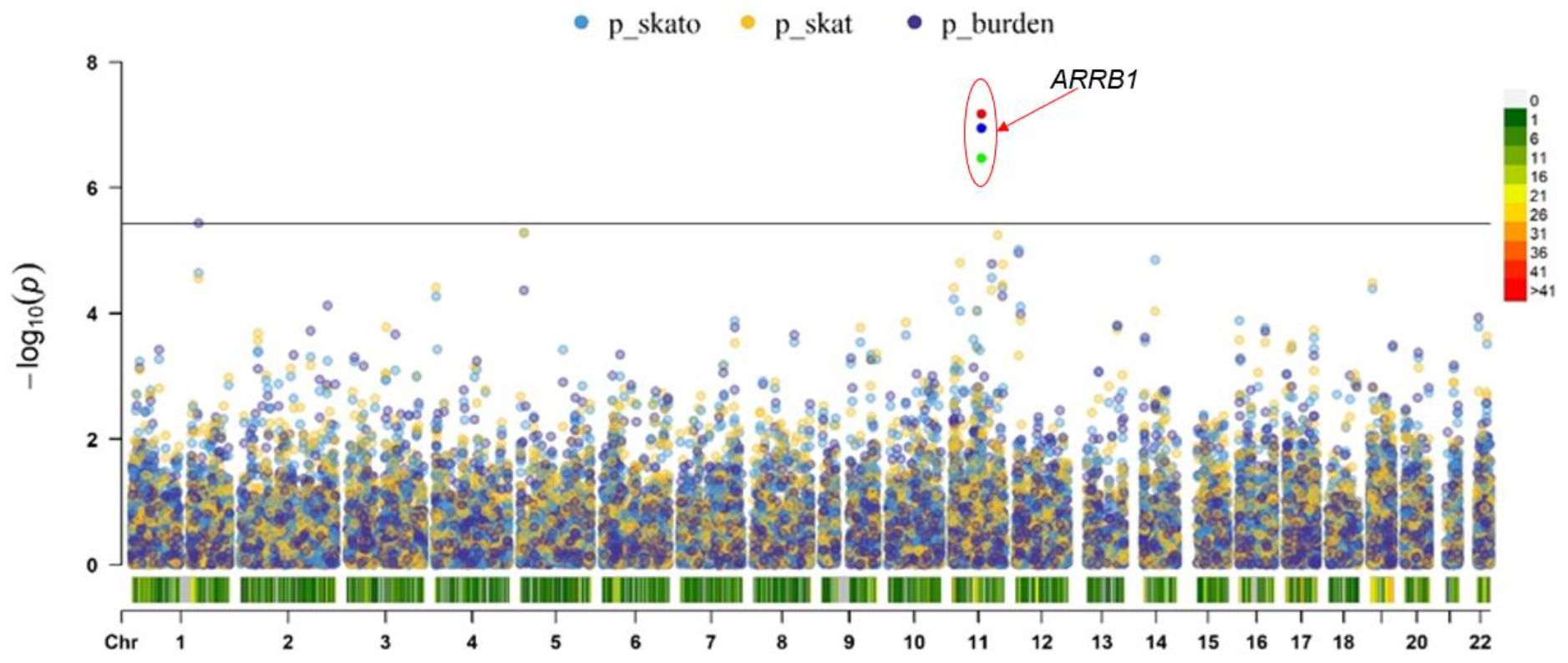
Gene-level association of low frequency and rare coding variants (MAF < 5%) using the burden, SKAT and SKAT-O association tests. Results of association with HbA1c reduction are shown for 13, 654 genes, adjusting for baseline HbA1c, age, sex, baseline BMI, and first 3 principal components. Genes with P < 3.66 × 10 ^-06^ are annotated.

Four variants, rs140226575G>A (Thr370Met, MAF = 2.2%), rs78979036G>A (Thr275Ile, MAF = 0.06%), rs58428187T>C (Ile158Val, MAF = 0.03%) and rs78052828T>C (Gly411Ser, MAF = 0.08%), contributed to the observed association. Compared to individuals without the risk alleles, carriers of one or more rare alleles showed greater reduction in HbA1c (1.09% ± 0.014 [11.9 mmol/mol ± 0.2] Vs 1.29% ± 0.064 [14.04 mmol/mol ± 0.7], p<0.001) (Figure 3). This association was mainly driven by a low frequency coding variant, rs140226575 (Thr370Met), minor allele frequency (MAF) = 2.2%, beta per allele = 0.25% ± 0.06 [2.7 mmol/mol ± 0.7], p=5.2×10^−06^. This is further replicated in the recently acquired data using the GLP-1RA arm of the HARMONY outcomes trial data^21^ (beta per allele = 0.14% ± 0.04 [1.5 mmol/mol ± 0.4], p=2.3×10^−04^). The frequency of rs140226575G>A (Thr370Met) is high in Hispanics (MAF 6%) and American Indians or Alaskan natives (MAF 11%), compared to White Europeans (MAF 0.5%), We therefore performed race stratified analysis and this revealed a consistent association between rs140226575G>A (Thr370Met) and glycaemic response to GLP-1RA with per A allele effect size of 0.18% ± 0.05 [2 mmol/mol ± 0.5], 0.17% ± 0.05 [1.8 mmol/mol ± 0.5], 0.11% ± 0.05 [1.2 mmol/mol ± 0.5] in White Europeans, Hispanics and American Indians or Native Alaskans, respectively (Supplementary Figure 4). Other suggestive gene-based results (p<1×10^−04^) in the overall meta-analysis are shown in Supplementary Table 5.

**Figure 3:**
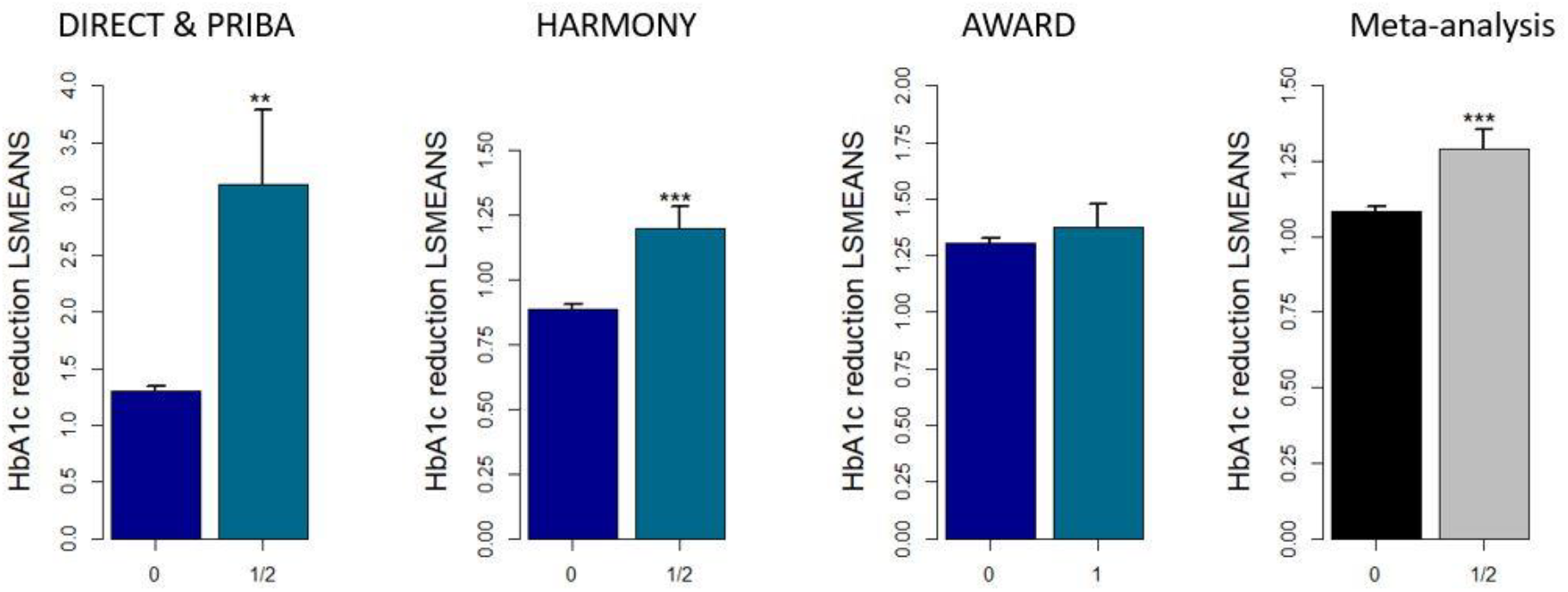
Bar plot of HbA1c post GLP-1RA therapy stratified by *ARRB1* variants (Thr370Met, Thr275Ile, Ile158Val, Gly411Ser). 00: wild type at all the SNPs and 1/2: those who carry one or more variant alleles at *ARRB1*.

### Genetic variants in the GLP-1R and ARRB1 are not associated with glycaemic response to metformin, sulphonylureas, pioglitazone, placebo or insulin treatment

To assess whether genetic variants in the *GLP-1R* and *ARRB1* that are associated with glycaemic response to GLP-1RA mark a general ability to respond to any other glucose lowering drugs, we examined association between the above variants and HbA1c reduction after treatment with metformin (n = 11, 933) or sulphonylureas (n = 5479). In addition, we analyzed comparator arms from the HARMONY phase 3 trials for pioglitazone (n = 191), placebo (n = 315), glimepiride (n = 207), or insulin (n=187). Neither the *GLP-1R* nor *ARRB1* variants were associated with glycaemic response to any of the non GLP-1RA treatment drugs including placebo (Supplementary Table 6). This data suggests a specific role of these variants on glycaemic response to GLP-1RA, rather than a general effect.

### Genetic variants in the GLP-1R and ARRB1 are not associated with the weight loss effect of GLP-1RA

Given treatment with GLP-1RA was associated with weight loss, we evaluated association between variants in the *GLP-1R* (Gly168Ser, Pro7Leu) and *ARRB1* with GLP-1RA related weight loss as a sensitivity analysis using data from the HARMONY phase 3 trials. None of the variants showed an association with weight loss (Supplementary Table 7).

### Genetic risk score derived from the GLP-1R and ARRB1 genes identify 4.2% of the population that had a clinically relevant greater response to GLP-1RAs

To evaluate the clinical relevance of *GLP-1R* and *ARRB1* in glycaemic response to GLP-1RA treatment, we categorized individuals based on their genotype at *GLP1-R* (Gly168Ser) and *ARRB1* (Thr370Met, Thr275Ile, Ile158Val, Gly411Ser) by aligning the alleles with the same direction of effect together as shown in Figure 4. The 4.2% of the population who carry one or more variant alleles in *ARRB1* had a mean (SEM) HbA1c reduction of 1.3% ± 0.1 [14.2 ± 0.8 mmol/mol] in response to GLP-1RA treatment. In contrast, 42.7% of the population who carry no variant allele at *ARRB1* and one or more Ser allele at position 168 of the *GLP-1R* had a mean (SEM) HbA1c reduction of 1% ± 0.02 [11 ± 0.3 mmol/mol], a difference of 0.3% [3.2 mmol/mol] (p < 0.001). Those who had no variant allele at *ARRB1* and no Ser allele at *GLP-1R* had a mean (SEM) HbA1c reduction of 1.1% ± 0.02 [12.4 ± 0.2 mmol/mol] (Figure 4).

**Figure 4:**
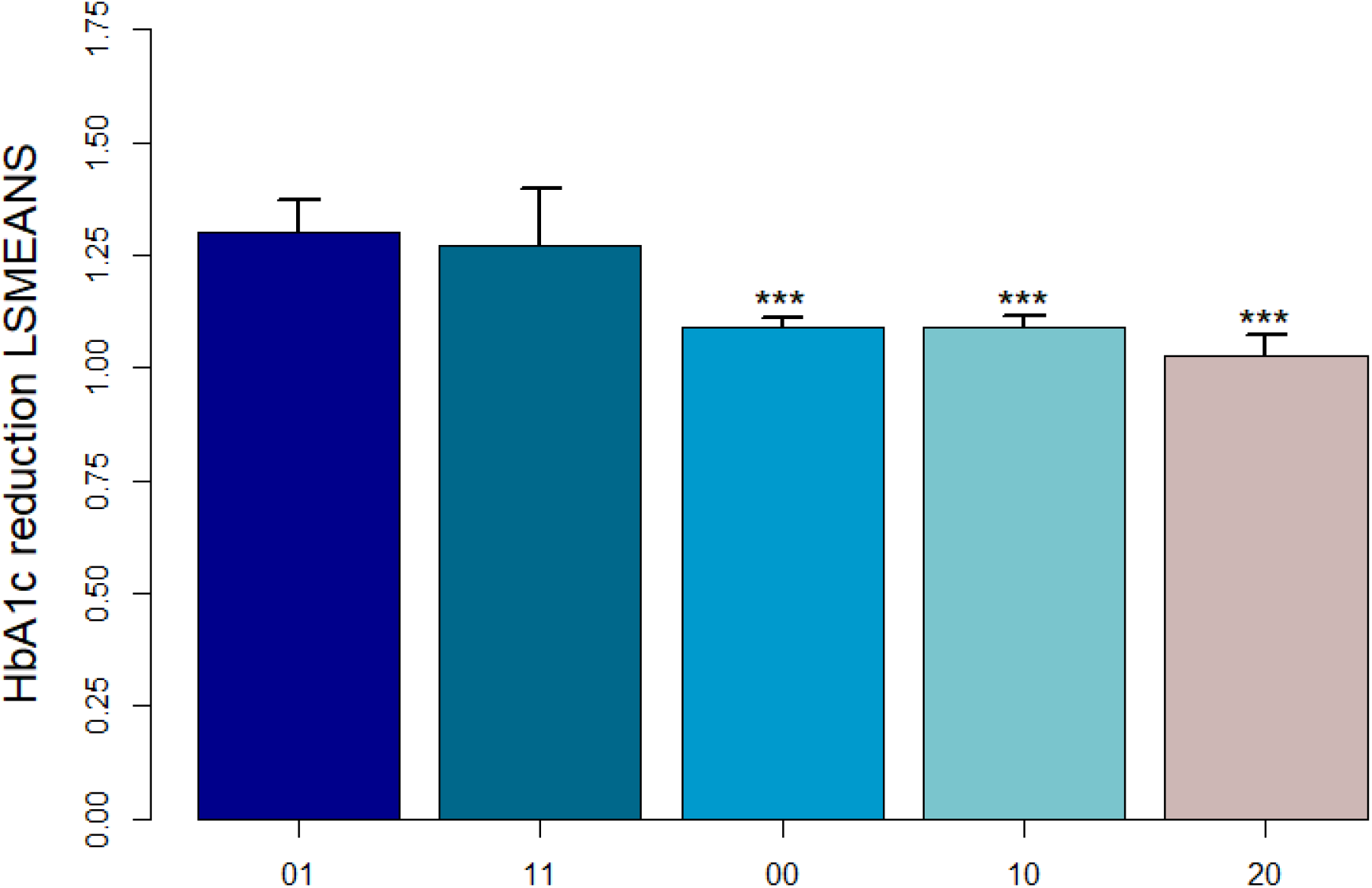
Bar plot of HbA1c post GLP-1RA therapy stratified by the genetic risk score from *GLP-1R* (Gly168Ser) and *ARRB1* SNPs (Thr370Met, Thr275Ile, Ile158Val, Gly411Ser). Bars represent mean HbA1c reduction and error bars represent standard error around the mean., ***p < 0.001 01: wild type at *GLP1R* and 1 or more variant alleles at any of the four *ARRB1* SNPs, 11: heterozygous at *GLP1R* and 1 or more variant alleles at *ARRB1*, 00: wild type at both *GLP1R* and *ARRB1*, 10: heterozygous at *GLP1R* and wild type at *ARRB1*, 20: two variant alleles at *GLP1R* and wild type at *ARRB1*.

## Discussion

We report an association of a common protein coding variant, rs6923761 (Gly168Ser), in the *GLP-1R* and low frequency and rare variants in the *ARRB1* genes with HbA1c reduction after treatment with GLP-1RA in 4,571 subjects across prospective observational and clinical trial data sets. A genetic risk score derived from these two genes identified 4.2% of the population who had a ∼30% greater reduction in HbA1c than the 42.7% of the population with the worse response.

### GLP-1 Receptor and GLP-1RA response

Our findings for the Gly168Ser, a *GLP-1R* variant, for GLP-1RA are supported by previous studies that resulted in this variant being selected as a candidate. In a small observational study this variant has previously been reported to be associated with glycaemic response to DPP4 inhibitors^11^. In addition, Gly168Ser was shown to be nominally associated with GLP-1 induced insulin secretion in healthy individuals studied by hyperglycaemic clamp, with carriers of the Ser allele showing ∼15% reduction in insulin secretion compared to homozygous carriers of the parent Gly allele^14^. In transfected Chinese hamster cell lines, the 168ser variant has previously been associated with reduced cell surface expression of the GLP-1R and reduced intracellular calcium mobilization following GLP-1 stimulation^22^. Interestingly, in the GWAS of GLP-1RA response, one of the variants suggestive of association was rs2268640, which is a cis eQTL for the *GLP-1R* in the pancreas.^23^ G-allele carriers had a -1.1 ± 0.22 mmol/mol worse response to GLP-1RA (2.52×10^−07^); the G allele is associated with reduced expression of *GLP-1R* in the pancreas (β = -0.19, p = 9.7×10^−04^)^24^ explaining the worse response (Supplementary Figure 3).

### Beta-Arrestin 1 and GLP-1RA response

Our finding that *ARRB1* variants are associated with greater glycaemic response to GLP-1RA followed a hypothesis free genome wide analysis. The beta-arrestins represent strong biological candidates for GLP-1RA response, as they have a key role in canonical G protein-coupled receptor (GPCR) internalization, with beta-arrestin recruitment targeting GPCR internalization via clathrin mediated mechanisms.^25,26^ However, there exists conflicting evidence for their role in controlling trafficking of incretin receptors.^27-31^ Depletion of beta-arrestins is reported to have little impact on agonist stimulated GLP-1R internalization.^28,31,32^ On the other hand, increased GLP-1R endocytosis has been reported after enhancing β-arrestin-2 action by the overexpression of G protein receptor kinase 5.^29^ Recently, selective reduction of β-arrestin-2 recruitment was also shown to reduce GLP-1R endocytosis and increased insulin secretion over a prolonged stimulation period.^30^ β-arrestins (both β-arrestin 1 and β-arrestin 2) recruitment to GLP-1R during acute and sustained agonist exposure has been shown to have opposite effect on insulin secretion.^33-36^ While beta-arrestins seemed to play important role for acute stimulation of insulin release, ß-cell knockout enhances insulin secretion.^33-36^ Our human pharmacogenetic studies resolve much of this uncertainty as they show convincingly that beta-arrestin 1 clearly has a role in glycaemic response to GLP-1RA, but not weight response to GLP-1RA pointing to a likely beta-cell mechanism. We show the 370Met allele associated with greater HbA1c reduction. Substitution of Thr370 by 370Met has been predicted to have possibly damaging effect on protein function.^37^ A reduction in β-arrestin 1 recruitment was shown to retain GLP-1R at the plasma membrane and thus produce greater long-term insulin release.^31^

We did find differences in HbA1c reduction by *ARRB1* genotype across cohorts. *ARRB1* variants had greater HbA1c reduction in users of exenatide and liraglutide (DIRECT & PRIBA) followed by albiglutide (HARMONY) and dulaglutide (AWARD) (Figure 3). This could potentially be because different GLP-1 agonists engage different signaling pathways, otherwise termed as biased agonism. Current evidence of bias between cAMP and β-arrestin recruitment on approved GLP-1RA is inconclusive.^38-41^ While this needs further investigation, G protein-biased GLP-1 agonists appear to achieve enhanced insulin release and thus greater glucose lowering efficacy by avoiding GLP-1 receptor desensitization and down-regulation, partly via reduced β-arrestin recruitment.^42-45^

### Genetic variants alter glycaemic response to GLP-1RA independent of weight change

In this study, none of the variants were significantly associated with weight loss consistent with the genetic variation altering pancreatic beta-cell response to GLP-1RA but not the gastric or hypothalamic actions of this drug class. Clinical trials and observational studies have shown a robust association of GLP-1RA treatment with weight loss in diabetic and non-diabetic subjects.^46^ Consistent with this, we see a mean 3.27kg weight loss in our study population. However, interindividual variation in weight reduction to GLP-1RA has been previously reported^47^. Gly168Ser and Pro7Leu have been investigated in relation to GLP-1RA induced weight loss. Some but not all studies have reported association of the Gly168Ser variant in *GLP-1R* with weight response to GLP-1RA treatment in T2D subjects.^48^ In a pilot study involving obese women with polycystic ovary syndrome, the Pro7Leu has also been associated with weight lowering potential of liraglutide.^12^

### Other potential genetic loci associated with GLP-1RA response

Whilst not genome wide significant there were a number of loci of potential interest in the GWAS and gene burden tests. The meta-GWAS yielded some variants with p<1×10^−05^ (Supplementary Table 4), including the *GLP-1R* eQTL SNP rs2268640 discussed earlier. The A allele at rs61800555 was associated with a -0.14% ± 0.03 [1.5 mmol/mol ± 0.4], reduction in HbA1c response to GLP-1RA (p = 2.51×10^−07^); this variant is in the intron of *LMX1A. LMX1A* encodes a transcription factor shown to act as a positive regulator of insulin gene transcription (Entrez Gene: LMX1A). In the gene burden tests, beyond *ARRB1, TAS2R1* was also above the GWAS significant threshold in the gene-based analysis (p-skato = 5.17×10^−06^). *TAS2R1* encodes a member of a family of candidate taste receptors that are members of the GPCR expressed in mucous epithelial cells of the tongue, colon, stomach and upper respiratory tract.^49-51^ Studies have shown that activation of *TAS2R1* can elicit stimulation of GLP-1.^52^ Expansion of the sample size and independent replication is required before we can be confident that these signals are definitely associated with GLP-1RA treatment response.

### Ethnicity and GLP-1RA response

The frequency of the minor allele in rs140226575G>A (Thr370Met), a variant that largely drives the *ARRB1* signal, varies by ethnicity with a frequency of 0.05%, 6% and 11% in White Europeans, Hispanics, and American Indians/Alaskan Natives, respectively and is almost monomorphic in populations of African and Asian descent. However, the effect size for associations between rs140226575 and GLP-1RA response was similar across ethnicities – which will result in greater population effect of this variant on GLP-1RA response among the Hispanics and American Indians/Alaskan Natives. Given this could have greater importance for clinical care among individuals of specific ancestry, there is a need to undertake well powered, diversity focused, multi-ethnic studies.

### Clinical translation of GLP-1RA pharmacogenomics

For treatment of common complex diseases like T2D, it is highly unlikely that genetic variants alone will be used to dictate treatment choice. This is in contrast to monogenic diabetes. We know that drug response is variable and influenced by many factors, and that treatment decisions are based on more than glucose levels. However, there is often treatment equipoise, with more than one treatment being suitable. We describe ∼5% of the population who have low frequency variants in *ARRB1* who respond 30% better (absolute benefit of 3.2mmol/mol) to GLP-1RA than the 43% of the population who have normal *ARRB1*, but a *GLP-1R* variant. Given that the average rate of glycaemic deterioration is 1mmol/mol/year^53^ these 5% will have 3-years longer before treatment failure, with the potential to use GLP-1RA earlier in their treatment regime. Clinically, GLP-1RA have benefits beyond glucose lowering – in particular, reduction in cardiovascular morbidity and mortality.^55^ It would be important to explore whether the genetic variants we have identified to alter glycaemic response to GLP-1RA also impact on cardiovascular risk, and to undertake genome wide studies on CV outcomes with GLP-1RA. Currently the data available are not sufficient to model these outcomes, especially for the rare variants we have identified in *ARRB1*.

### Conclusions

We have undertaken the first large-scale candidate based and genome wide analysis of glycaemic response to GLP-1RA and have identified that variants in genes encoding the GLP-1 receptor and beta-arrestin 1 alter glycaemic response to this drug class. This has provided novel insight into GLP-1 signalling in the pancreatic beta-cell. We establish that a coding variant previously shown to reduce GLP-1R membrane expression and GLP-1 stimulated insulin secretion in beta-cells reduces GLP-1RA response. We also provide clarity that in humans that beta-arrestin 1 does play a role in glycaemic response to GLP-1RA. Finally, we maintain that the genetic effect sizes identified can add to clinical data to identify a subgroup of patients who respond particularly well to GLP-1RA supporting early use of these drugs for those individuals, especially in Hispanic and American Indian populations.

## Data Availability

All data produced in the present study are available upon reasonable request to the authors

## Contributors

Conception and design of the study: ERP, AYD, AGJ, TJM, LMH, AM, IP, HR, MW; Data analysis: AYD, PB, LL, SW, KZ, MH, SH; data collection and genotyping: NRR, AM, IF, AGJ, TJM; AYD, and ERP wrote and prepared the manuscript. All authors were involved in reading and editing the manuscript. AYD and ERP are guarantors of this work.

## Data sharing

Summary-level data that underlie the results reported in this article will be available upon request to the corresponding author.

## Acknowledgements

We would like to thank the MetGen and MetGen Plus consortiums for provision of summary level data for metformin and sulphonylurea response.

