## Supplementary material for "Pharmacogenomics of GLP-1 Receptor Agonists: A genome wide analysis of observational data and large randomized controlled trials"

**Supplementary documents**

***Cohort descriptions***

DIRECT

Three groups of subjects with type 2 diabetes from four IMI-DIRECT (DIabetes REsearCh on patient straTification) participating centres have been included in this cohort. The first group were recruited just before starting a GLP-1RA and followed up for 6 months. The second group were recruited after they had GLP-1RA treatment between 6 and 24 months and were still on treatment at the time of assessment. The third group were all patients who had ever been treated with a GLP-1RA where they had at least 4 months of GLP-1RA treatment and an HbA1c measurement within the 6 weeks prior to starting the GLP-1RA, and within 6 months (± 2 months) after starting the GLP-1RA in order to define HbA1c reduction. The inclusion criteria for all the groups were a) baseline HbA1c ≥ 7.5% (58mmol/mol) and HbA1c < 12% (108 mmol/mol), b) White European, c) Age ≥ 18 years and < 80 years. Approval for the study protocol was obtained from each of the regional research ethics review boards separately (Lund, Sweden: 20130312105459927, Copenhagen, Denmark: H-1-2012-166 and H-1-2012-100, Amsterdam, Netherlands: NL40099.029.12, Newcastle, Dundee and Exeter, UK: 12/NE/0132) and all participants provided written informed consent at enrolment. The research conformed to the ethical principles for medical research involving human participants outlined in the declaration of Helsinki.

PRIBA

The PRIBA (Predicting Response to Incretin Based Agents in Type 2 Diabetes) study recruited subjects with type 2 diabetes commencing GLP-1RA as part of routine care prospectively between April 2011 and October 2013 with baseline HbA1c ≥ 7.5% (58 mmol/mol) and estimated glomerular filtration rate (eGFR) > 30 mL/min/1.73m$^2$. Participants were assessed (including HBA1c) immediately prior to commencing treatment and after 3 and 6 months on therapy. All the participants signed informed consent and the Southwest National Research Ethics committee approved the study. Further study details, and details of previous publications, are available through https://clinicaltrials.gov/ct2/show/NCT01503112

GoDARTS

GoDARTS (Genetics of Diabetes Audit and Research in Tayside Scotland) is a longitudinal cohort study established to study the genetics of type 2 diabetes. Over 18,000 participants were enrolled between December 1998 and August 2012, of whom half of them are diagnosed with type 2 diabetes and the remaining age and sex matched non-diabetic controls identified from general practice records in Tayside, Scotland.[1] Comprehensive electronic medical records history dating back to 1990 including anthropometric, clinical, prescription and biochemistry is available for each participant through a unique anonymised community health index number provided by the Health Informatics Centre (HIC) in partnership with the University of Dundee and the National Health Service (NHS).

From 659 incident GLP-1RA users in the GoDARTS cohort, we identified a study sample of 315 patients who had been started on liraglutide or exenatide as their second-line (added to metformin or sulphonylurea monotherapy) or third-line (added to metformin and/or sulphonylurea and/or thiazolidinediones, dual therapy) treatment according to the NHS guideline in Scotland. All the patients in the study had complete data with respect to age, gender, antidiabetic treatment history, regular HbA1c measurements and genotype. They all had a baseline HbA1c higher than 7% (53 mmol/mol). They were on stable treatment for 6 months after GLP-1RA was initiated (the index date), which meant they did not start or stop another antidiabetic drug within the study period. They were not treated with insulin before or during the study period. The study was approved by the Tayside Regional Ethics Committee and informed consent was obtained from all subjects.

PROMASTER

The PROMASTER (PROspective Cohort MRC ABPI STratification and Extreme Response Mechanism in Diabetes) is a longitudinal study designed to examine extreme responders to second- and third-line type 2 diabetes therapies using a prospective approach. All the participants were clinically diagnosed with type 2 diabetes aged between 18 and 90 years, had HbA1c > 7% (53 mmol/mol). All the participants signed informed consent and the Southwest National Research Ethics committee approved the study. Data from 79 participants treated with GLP-1RA as part of their diabetes care were included for this meta-analysis. Further study information is available through https://www.clinicaltrials.gov/ct2/show/NCT02105792.

The HARMONY trials

The HARMONY program for albiglutide includes eight phase III clinical trials designed to evaluate the efficacy and safety of albiglutide in patients with type 2 diabetes inadequately controlled on lifestyle and/or a combination of other glucose lowering drugs. Data from the

GLP-1RA (Albiglutide) arm of seven of these trials is included in the current study.[2-8] Supplementary Table 1 shows the different studies included with numbers in each arm with background medication and duration for primary end points. Each study included male and nonpregnant, nonlactating female participants of 18 years of age or older, with a history of type 2 diabetes diagnosis currently on lifestyle and/or other hypoglycaemic agents but experiencing inadequate glycaemic control. Patients were required to have baseline HbA1c between 7.0 and 10.0% (53–86 mmol/mol), BMI between 20 and 45 kg/m$^2$

Important exclusion criteria included a history of cancer (except non-melanoma skin cancers) not in remission for 3 years), treated diabetic gastroparesis, current symptomatic biliary disease or history of pancreatitis, significant gastrointestinal surgery, or recent significant cardiovascular or cerebrovascular events and history or family history of medullary carcinoma or multiple endocrine neoplasia type 2. Other study specific inclusion/exclusion criterion are available in the respective publications.[2-8]. For the HARMONY studies, approval is provided by an independent committee who manage access to clinical trials (https://www.clinicalstudydatarequest.com/). All study participants have provided informed consent.

For this study, participants randomized to the GLP-1RA arm who have genotypic and complete clinical data are included. Glycaemic response to GLP-1RA was modelled as the quantitative phenotype of HbA1c reduction between baseline HbA1c and treatment HbA1c. Baseline HbA1c was the HbA1c measure at randomization. The treatment HbA1c was the closest HbA1c measure to 6 months after initiation of GLP-1RA (between 3 and 9 months).

**Supplementary Table 1: HARMONY phase 3 trials included in the current analysis**

| **Study** | **Treatment arm at randomization (n)** | **Comparator arm at randomization (n)** | **Background medication** | **Duration of primary end point (weeks)** | **Genotyped** |
|---|---|---|---|---|---|
| HARMONY 1 | Albiglutide (155) | Placebo (155) | Pioglitazone and/or metformin | 52 | 244 |
| HARMONY 2 | Albiglutide (204) | Placebo (105) | Diet and exercise | 52 | 234 |
| HARMONY 3 | Albiglutide (302) | Placebo (101)/Sitagliptin (302)/Glimepiride (307) | Metformin | 104 | 730 |
| HARMONY 4 | Albiglutide (504) | Insulin (241) | Metformin and/or sulfonylurea | 52 | 584 |
| HARMONY 5 | Albiglutide (281) | Placebo (116)/Pioglitazone (288) | Metformin and glimepiride | 52 | 490 |

| | | | | | |
|---|---|---|---|---|---|
| HARMONY 7 | Albiglutide (422) | Liraglutide (419) | Any oral medication | 32 | 638 |
| HARMONY 8 | Albiglutide (254) | Sitagliptin (253) | Diet and or exercise and/or any oral medication | 26 | 290 |
| Total | 2, 122 | 2, 287 | | | 3210 |

AWARD trials:

The **A**ssessment of **W**eekly **A**dministR**a**tion of LY2189265 (dulaglutide) in **D**iabetes (AWARD) is a phase 3 clinical study program that compared GLP1-RA dulaglutide (0.75 mg and/or 1.5 mg) to a variety of common antihyperglycemic medications. Studies were designed to assess efficacy, safety, and patient reported outcomes in patients across different stages of the T2D treatment. Data from the dulaglutide arms along with two other active GLP1-RA comparators (exenatide and liraglutide) from five trials were included in this study[9-13]. Supplementary Table 2 shows different studies included with numbers in each arm, background medications and duration for primary end points. The primary outcome in AWARD studies was HbA1c change from baseline to the primary endpoint were mostly 26-52 weeks depending on the study. To achieve consistency in our analyses, we used 26 or 24 weeks HbA1c change from baseline as our primary endpoint. The participants with full consent and available genotypic and clinical data were only included in this analysis. The protocol for this study was approved by an independent committee who manage access to clinical trials (https://www.clinicalstudydatarequest.com/). All study participants have provided informed consent.

**Supplementary Table 2: AWARD phase 3 trials included in the current analysis**

| Study | at randomization (n) | Comparator(s) (dose) - (n) | Background medication | Duration of primary end point (weeks) | Genotyped (n) | Analyzed (n=1,562) |
|---|---|---|---|---|---|---|
| AWARD-1 (GBDA)§ | Dulaglutide (498) | Exenatide (10µg BID) - (249) Placebo | Pioglitazone (≥30 mg) Metformin (≥1500 mg) | 26 | Dulaglutide (341) Exenatide (166) | 501 |
| AWARD-2 (GBDB)§ | Dulaglutide (446) | Insulin Glargine (titrated to target) | Glimepiride (≥4 mg) Metformin (≥1500 mg) | 52 | Dulaglutide (187) | 185 |
| AWARD-5 (GBCF)§ | Dulaglutide (606) | Sitagliptin (100 mg QD) Placebo | Metformin (≥1500 mg) | 52 | Dulaglutide (179) | 178 |
| AWARD-6 (GBDE)§ | Dulaglutide (299) | Liraglutide (1.8 mg QD) - (300) | Metformin (≥1500 mg) | 26 | Dulaglutide (243) Liraglutide (233) | 476 |

| | | | | | | |
|---|---|---|---|---|---|---|
| AWARD-8 (GBDG) ¶ | Dulaglutide (239) | Placebo | Sulfonylurea (at least 50% of maximum tolerated dose) | 24 | Dulaglutide (225) | 222 |

§ Indicates that genetic analyses for HbA1c reduction were conducted at 26 weeks change from baseline (CFBL); ¶ Indicates that genetic analyses for HbA1c reduction were conducted at 24 weeks change from baseline (CFBL)

HARMONY outcome trial

The HARMONY outcome was a double-blind, randomised, placebo-controlled trial designed to evaluate the cardiovascular benefit of Albiglutide in patients with type 2 diabetes[14]. Each study included men and women aged 40 years or older with established disease of the coronary, cerebrovascular, or peripheral arterial circulation who had a glycated haemoglobin concentration of more than 7·0% (53 mmol/mol). Individuals with estimated glomerular filtration rate (eGFR) of less than 30 mL/min per 1·73 $m^2$, severe gastroparesis, previous pancreatitis or substantial risk factors for pancreatitis, a personal or family history of medullary carcinoma of the thyroid or multiple endocrine neoplasia type 2, a history of pancreatic neuroendocrine tumours, or current use of a GLP-1A were excluded[14]. Patients had study visits every four months - anthropometric and biochemical measures were collected. The protocol for this study was approved by an independent committee who manage access to clinical trials (https://www.clinicalstudydatarequest.com/). All study participants have provided informed consent.

For this study, participants randomized to the GLP-1RA arm with genotype data for rs140226575 and complete clinical data are included. Glycaemic response to GLP-1RA was modelled as the quantitative phenotype of HbA1c reduction between baseline HbA1c and treatment HbA1c. Baseline HbA1c was the HbA1c measure at randomization. The treatment HbA1c was the HbA1c measure at 8 months after initiation of GLP-1RA.

***Genotyping and imputation***

Genotypes for the DIRECT, PRIBA and PROMASTER studies were generated using the Illumina Human Core Exome chip v1.1 (HCE24 v1.1). Genotype calling for both common and low-frequency variants was performed using the GenCall algorithm in the GenomeStudio software supplied by Illumina. Data were subjected to a series of standard quality control analyses in order to highlight poorly performing genetic markers and samples prior to imputation. Individuals that had a call rate lower than 97% were excluded. The heterozygosity rate per sample was calculated using the formula (number of non-missing genotypes N (NM) - Number of homozygous genotypes O (Hom)) / N (NM). Cut-off for exclusion of outliers was 4 standard deviations from the mean heterozygosity rate. Gender check was ascertained to detect discrepancies between phenotypic and genotypic sex. Individuals with discordant sex information were removed. In order to avoid bias from duplicates and related individuals, estimates of pairwise identity by decent (IBD) were generated. Among the related samples, IBD > 0.2, the one with the lowest call rate was excluded. Each dataset was then imputed to the 1000 Genomes Phase 3 CEU reference panel with ShapeIt (v2.r790)[15] and Impute2 (v2.3.2).[16]

For the GoDARTs data, a single time point blood sample was collected from each participant for DNA extraction and genotyping. Each of the Illumina Omni-express (Illumina, San Diego, USA) and the Affymetrix 6.0 SNP (Affymetrix, Santa Clara, USA) genotyping arrays were used to genotype participants with T2D. After standard quality control of the genotypic data, haplotypes were estimated using ShapeIt (v2.r790)[15] and imputation of the missing genotypes was performed using the 1000 Genomes Phase 3 CEU reference panel with Impute2 (v2.3.2).[16] Missing alleles were imputed by running a forward-backward algorithm with a certain probability.

HARMONY phase 3: Samples from consented participants were genotyped with the Affymetrix Axiom Array with custom content (GSKBB1_v1). Genotypes were reported on the forward strand of the GRCH37/hg19 assembly. Participants with call rates < 96% and discrepancies in reported sex were removed. Samples with extra heterozygosity (more than 4 standard deviation away from the mean) or correlated with another sample (identity by descent [IBD]>0.2) were filtered out. Variants that are monomorphic or with call rates < 95% were removed. Autosomes were imputed to the 1000G panel using Michigan Imputation Server and converted to hard calls using default PLINK threshold settings. Non-biallelic SNPs were filtered out and SNPs with imputation quality ($R^2$) less than 0.3. SNPs with genotyping rates < 0.95, Hardy Weinberg Equilibrium P-value less than $10x10^{-6}$ were excluded. Individuals with call rates less than 0.96 were excluded. A GWAS was performed using SNPTEST[17] including sex, baseline HbA1c, baseline BMI, duration of diabetes, study, and the top 10 principal components as covariates.

AWARD phase 3

Human genomic DNA from 5 dulaglutide trials (AWARD 1, 2, 5, 6 and 8) enrolled participants was extracted using peripheral whole blood. Participant who consented for genetic analyses were only included in the study. Genome-wide data was generated using the Illumina's HumanOmni-5 exome array (Santa Clara, CA, US) and standard quality control metrics of genome-wide association study were applied. Samples with call rate < 95% across all variants and discrepancies in reported sex were removed. Identity-by-state (IBS) score for all possible pairs of subjects was calculated and subjects with unusually high IBS scores were excluded from the analyses. Variants that were monomorphic or with call rates < 95% were removed. Samples that passed QC were later used as an input to perform genome-wide imputation. Chromosomal phasing was performed with Beagle v.4.1 using GRCh37/hg19 map reference file, and later genome-wide data imputation was performed using Minimac3 algorithm (https://genome.sph.umich.edu/wiki/Minimac3). The 1,000 Genomes Project phase 3 data from various ethnic groups (ftp://ftp.1000genomes.ebi.ac.uk/vol1/ftp/phase3/data/) was used as the reference for the imputation. The imputation accuracy was evaluated using metrics generated by minimac3. The empirical correlation (Pearson correlation coefficient between Leave-One-Out dosages and known genotypes) was used to evaluate the imputation accuracy for genotyped variants. The average empirical correlation for variants that belong to different MAF bins was calculated to check the impact of MAF on imputation accuracy. For all downstream analyses, only biallelic autosomal variants were considered and optimal thresholds for Rsq were recommended given evaluation with various MAF ranges: an Rsq threshold of 0.3 is recommended for MAF > 1%; and threshold of 0.8 is recommended for MAF ≤ 1%.

Seven separate GWASs (5 dulaglutide, 1 liraglutide and 1 exenatide) were performed using a model that consist of HbA1c (change from baseline) as response variable, and genotype, baseline HbA1c and top 3 principal components as fixed effect variables, whereas study was used as random effect in the model. Treatment dose was used as a covariate if multiple doses were used in the study.

**Supplementary Table 3: Baseline characteristics of participants included in each cohort**

| Characteristics | DIRECT | PRIBA | GoDARTS | PROMASTER | HARMONY Phase 3 | AWARD | HARMONY outcomes |
|---|---|---|---|---|---|---|---|
| n | 365 | 471 | 323 | 79 | 1771 | 1562 | 3748 |
| Age (years) | 59.72 ± 9.95 | 55.99 ± 10.18 | 59.16 ± 8.82 | 54.61 ± 11.43 | 55.81 ± 10.08 | 55.93 ± 9.59 | 64.29 ± 8.64 |
| Duration of diabetes (years) | 12.73 ± 6.58 | 10.02 ± 6.56 | 10.64 ± 4.72 | - | 8.00 ± 6.11 | 8.07 ± 5.36 | 14.12 ± 8.49 |
| Sex (% women) | 148 (40.6%) | 216 (45.6%) | 138 (42.7%) | 35 (44.3%) | 830 (46.9%) | 773 (49.5%) | 1103 (29.4%) |
| Pre-treatment weight (kg) | 111.21 ± 22.08 | 114.47 ± 22.86 | 110.60 ± 22.04 | 106.12 ± 18.96 | 93.8 ± 20.51 | 92.21 ± 18.66 | 92.94 ± 19.86 |
| Pre-treatment BMI (kg/m$^2$) | 38.69 ± 6.94 | 39.77 ± 7.49 | 38.33± 6.71 | 36.99 ± 6.91 | 33.14 ± 5.62 | 32.77 ± 5.23 | 32.50 ± 5.98 |
| Pre-treatment HbA1c (DCCT-%) | 9.39 ± 1.20 | 9.76 ± 1.58 | 9.42 ± 1.39 | 9.85 ± 1.59 | 8.16 ± 0.67 | 8.14 ± 1.04 | 8.69 ± 1.41 |
| On-treatment weight (kg) | 107.14 ± 22.47 | 110.10 ± 21.78 | 112.85 ± 22.15 | 104.09 ± 20.13 | 92.98 ± 20.46 | 90.41 ± 18.81 | 91.87 ± 19.72 |
| On-treatment BMI (kg/m$^2$) | 37.25 ± 7.09 | 38.10 ± 7.05 | 39.07 ± 6.68 | 36.25 ± 6.94 | 32.84 ± 5.56 | 32.12 ± 5.29 | 32.13 ± 5.96 |
| On-treatment HbA1c (DCCT-%) | 8.19 ± 1.45 | 8.38 ± 1.59 | 8.67 ± 1.65 | 8.94 ± 1.82 | 7.26 ± 0.95 | 6.83 ± 0.99 | 7.40 ± 1.35 |
| Weight fall (kg)[#] | 4.00 [1.00-7.28] | 3.45 [0.90-6.90] | 4.45 [1.85-8.15] | 2.75 [0.28-4.45] | 0.84 [-1.0 – 2.40] | 1.6 [-0.05 - 4.00] | 0.80 [-1.00 – 2.80] |
| BMI fall (kg/m$^2$)[#] | 1.47 [0.38-2.59] | 1.17 [0.31-2.37] | 1.72 [0.67-2.86] | 0.88 [0.10-1.55] | 0.24 [-0.39-0.91] | 0.6 [-0.16 – 1.40] | 0.27 [-0.35 – 0.99] |
| HbA1c fall (DCCT-%) | 1.20 ±1.34 | 1.38 ±1.55 | 0.75 ± 1.71 | 0.89 ± 1.64 | 0.90 ± 0.89 | 1.31 ± 1.00 | 0.98 ± 1.33 |
| Ethnicity - White Europeans | 365 (100%) | 471 (100%) | 323 (100%) | 79 (100%) | 950 (54%) | 1151 (74%) | 2784 (74%) |
| Ethnicity - Hispanics | - | - | - | - | 386 (22%) | 63 (4%) | 546 (15%) |
| Ethnicity - American Indians/Alaska Native | - | - | - | - | 123 (7%) | 189 (12%) | 231 (6%) |
| Ethnicity - Others | - | - | - | - | 312 (17%) | 159 (10%) | 187 (5%) |

BMI, body mass index. [#] Median [IQR]

**Supplementary Table 4: Results for index variants at suggestive loci ($p<1\times10^{-5}$) associated with glycaemic response identified in a GWAS meta-analysis of GLP-1RA users with type 2 diabetes.**

| RSID | Chr | Position | EA | NEA | EAF | beta | se | Nearest gene | p.value | n_samples |
|---|---|---|---|---|---|---|---|---|---|---|
| rs61800555 | 1 | 165220705 | A | G | 0.195 | -0.142 | 0.025 | *LMX1A* | $2.51\times10^{-07}$ | 4462 |
| rs2268640 | 6 | 39050384 | A | G | 0.608 | 0.100 | 0.020 | *GLP-1R* | $2.52\times10^{-07}$ | 4462 |
| rs7687008 | 4 | 139268507 | C | T | 0.404 | 0.104 | 0.020 | *LINC00499* | $1.55\times10^{-06}$ | 4462 |
| rs10224036 | 7 | 77106630 | G | A | 0.190 | 0.133 | 0.026 | *PTPN12* | $1.98\times10^{-06}$ | 4462 |
| rs1969320 | 2 | 131027031 | G | C | 0.612 | 0.104 | 0.021 | *MTND1P29* | $3.52\times10^{-06}$ | 4150 |
| rs11072298 | 15 | 71854982 | C | A | 0.688 | -0.106 | 0.021 | *THSD4* | $5.23\times10^{-06}$ | 4462 |
| rs4986076 | 17 | 81027889 | A | G | 0.086 | 0.171 | 0.035 | *METRNL* | $5.31\times10^{-06}$ | 4463 |
| rs2048020 | 16 | 32375625 | T | G | 0.062 | 0.321 | 0.065 | *LOC105371191* | $5.88\times10^{-06}$ | 2406 |
| rs2298192 | 10 | 132915050 | A | G | 0.148 | -0.137 | 0.028 | *TCERG1L* | $6.64\times10^{-06}$ | 4462 |
| rs10561032 | 19 | 5742326 | G | T | 0.153 | 0.143 | 0.029 | *CATSPERD* | $7.18\times10^{-06}$ | 4150 |
| rs56354900 | 16 | 77451045 | T | A | 0.604 | -0.100 | 0.020 | *ADAMTS18* | $7.53\times10^{-06}$ | 4150 |
| rs11746176 | 5 | 78987399 | G | C | 0.208 | 0.119 | 0.024 | *CMYA5* | $7.54\times10^{-06}$ | 4462 |
| rs40182 | 5 | 1350397 | A | G | 0.382 | -0.099 | 0.020 | *CLPTM1L* | $7.76\times10^{-06}$ | 4462 |
| rs5767119 | 22 | 48758497 | T | C | 0.133 | 0.139 | 0.029 | *LOC105373081* | $8.98\times10^{-06}$ | 4462 |
| rs2286414 | 7 | 157475020 | T | C | 0.053 | 0.202 | 0.042 | *PTPRN2* | $9.19\times10^{-06}$ | 4463 |
| rs75941546 | 3 | 187866416 | A | G | 0.082 | 0.171 | 0.036 | *LPP* | $9.60\times10^{-06}$ | 4462 |

*EA: Effective allele, †NEA: Non-effective allele, EAF: Effective allele frequency, A negative beta implies that the effective allele is associated with reduced response to GLP-1RA.

**Supplementary Table 5: Genes most strongly associated with HbA1c reduction in a gene-based analysis, with $p < 1.0\times10^{-04}$.**

| gene | Extended gene name | p_burden | p_skat | p_skato | nsnps |
|---|---|---|---|---|---|
| *ARRB1* | Arrestin beta 1 | $1.12\times10^{-07}$ | $3.41\times10^{-07}$ | $6.72\times10^{-08}$ | 4 |
| *TAS2R1* | Taste receptor type 2-member 1 | $4.31\times10^{-05}$ | $5.27\times10^{-06}$ | $5.17\times10^{-06}$ | 3 |
| *NANOGNB* | NANOG Neighbor Homeobox | $1.09\times10^{-05}$ | $4.67\times10^{-04}$ | $9.65\times10^{-06}$ | 2 |
| *PYGL* | Glycogen Phosphorylase L | $2.81\times10^{-05}$ | $9.24\times10^{-05}$ | $1.42\times10^{-05}$ | 11 |
| *PRRX1* | Paired Related Homeobox 1 | $3.69\times10^{-06}$ | $2.82\times10^{-05}$ | $2.30\times10^{-05}$ | 3 |
| *TSPAN33* | Tetraspanin 33 | $5.75\times10^{-04}$ | $2.53\times10^{-05}$ | $2.33\times10^{-05}$ | 2 |
| *BIRC3* | Baculoviral IAP Repeat Containing 3 | $1.63\times10^{-05}$ | $4.27\times10^{-05}$ | $2.72\times10^{-05}$ | 7 |
| *KCNJ1* | Potassium Inwardly Rectifying Channel Subfamily J Member 1 | $5.29\times10^{-05}$ | $3.53\times10^{-05}$ | $3.89\times10^{-05}$ | 6 |
| *TNFSF14* | TNF Superfamily Member 14 | $1.49\times10^{-03}$ | $3.29\times10^{-05}$ | $4.01\times10^{-05}$ | 5 |
| *RNF4* | RING finger protein 4 | $1.28\times10^{-02}$ | $3.90\times10^{-05}$ | $5.36\times10^{-05}$ | 4 |
| *TRIM21* | Tripartite motif containing-21 | 0.59 | $3.91\times10^{-05}$ | $5.98\times10^{-05}$ | 9 |
| *GPRC5A* | G Protein-Coupled Receptor Class C Group 5 Member A | $1.03\times10^{-04}$ | $1.31\times10^{-04}$ | $7.89\times10^{-05}$ | 12 |
| *SLC6A5* | Solute carrier family 6 member 5 | $4.85\times10^{-03}$ | $1.56\times10^{-05}$ | $9.16\times10^{-05}$ | 9 |
| *EHD1* | EH domain-containing protein 1 | $8.16\times10^{-03}$ | $8.87\times10^{-05}$ | $9.24\times10^{-05}$ | 2 |

**Supplementary Table 6: Association of *GLP-1R* variants and *ARRB1* with HbA1c reduction after treatment with different glucose lowering drugs.**

| SNP | Beta ± SE | P |
|---|---|---|
| **Metformin (n = 11933)** | | |
| rs6923761G>A | **0.007 ± 0.012** | **0.54** |
| rs10305420C>T | 0.017 ± 0.013 | 0.20 |
| **Sulphonylureas (n = 5479)** | | |
| rs6923761G>A | -0.004 ± 0.021 | 0.87 |
| rs10305420C>T | -0.011 ± 0.024 | 0.65 |
| | **Placebo (315)** | |
| rs6923761G>A | 0.115 ± 0.098 | 0.24 |
| rs10305420C>T | 0.121 ± 0.086 | 0.16 |
| ARRB1 GRS | 0.135 ± 0.222 | 0.54 |
| **Pioglitazone (n = 191)** | | |
| rs6923761G>A | -0.115 ± 0.104 | 0.27 |
| rs10305420C>T | 0.059 ± 0.092 | 0.52 |
| ARRB1 GRS | 0.329 ± 0.255 | 0.20 |
| **Glimepiride (n = 207)** | | |
| rs6923761G>A | 0.068 ± 0.115 | 0.56 |
| rs10305420C>T | 0.052 ± 0.106 | 0.63 |
| ARRB1 GRS | -0.167 ± 0.277 | 0.55 |
| **Insulin (n = 187)** | | |
| rs6923761G>A | -0.078 ± 0.132 | 0.60 |
| rs10305420C>T | -0.002 ± 0.117 | 0.99 |
| *ARRB1* GRS | -1.017 ± 0.706 | 0.15 |

**Supplementary Table 7: Association of *GLP-1R* variants and *ARRB1* with BMI reduction after treatment with GLP-1RA.**

| SNP | Beta ± SE | P |
|---|---|---|
| rs6923761G>A | 0.022 ± 0.05 | 0.68 |
| rs10305420C>T | -0.06 ± 0.05 | 0.49 |
| ARRB1 GRS | 0.02 ± 0.13 | 0.88 |

*Supplementary Figures*

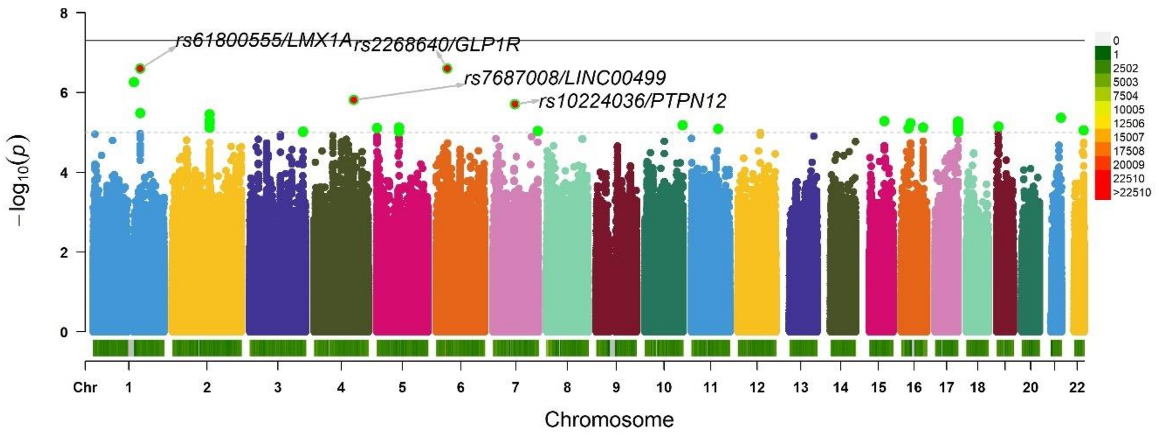


Supplementary Figure 1: Manhattan plot of single variant association results from a linear regression in the overall meta-analysis (n = 4,563). The–log10 p-values for each test are plotted against chromosomal position. A genome-wide significance threshold of 5 $\times 10^{-08}$ is indicated by the red horizontal bar.

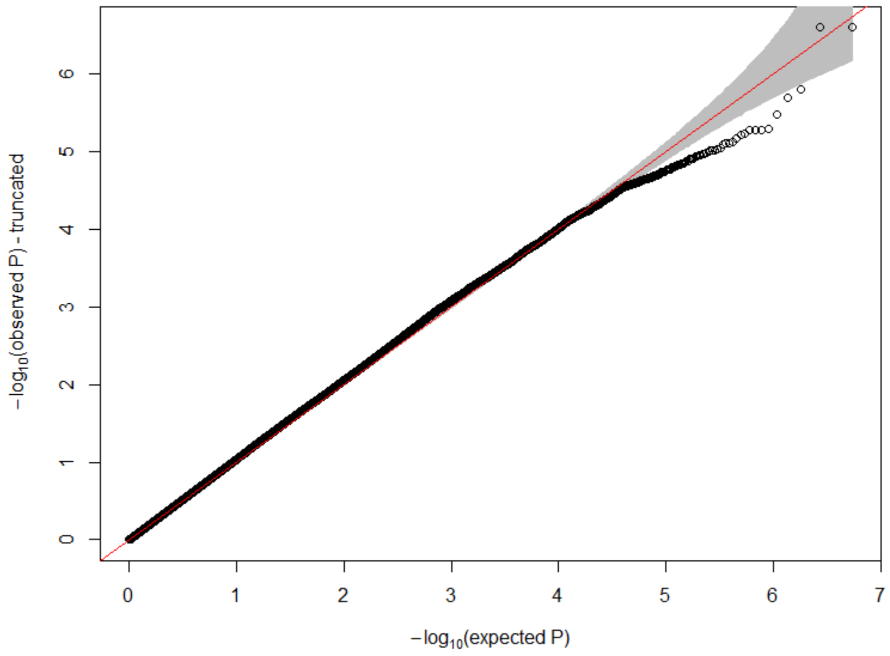


Supplementary Figure 2: The relationship between observed and expected p values (Q-Q plot) ($\lambda$ = 1.04).

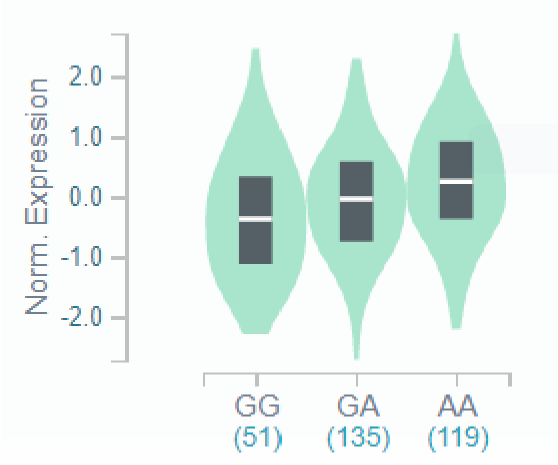


Supplementary Figure 3: eQTL of GLP-1R variant, rs2268640 on GLP-1R expression in the pancreas (Violin plot generated from the GTEx portal).

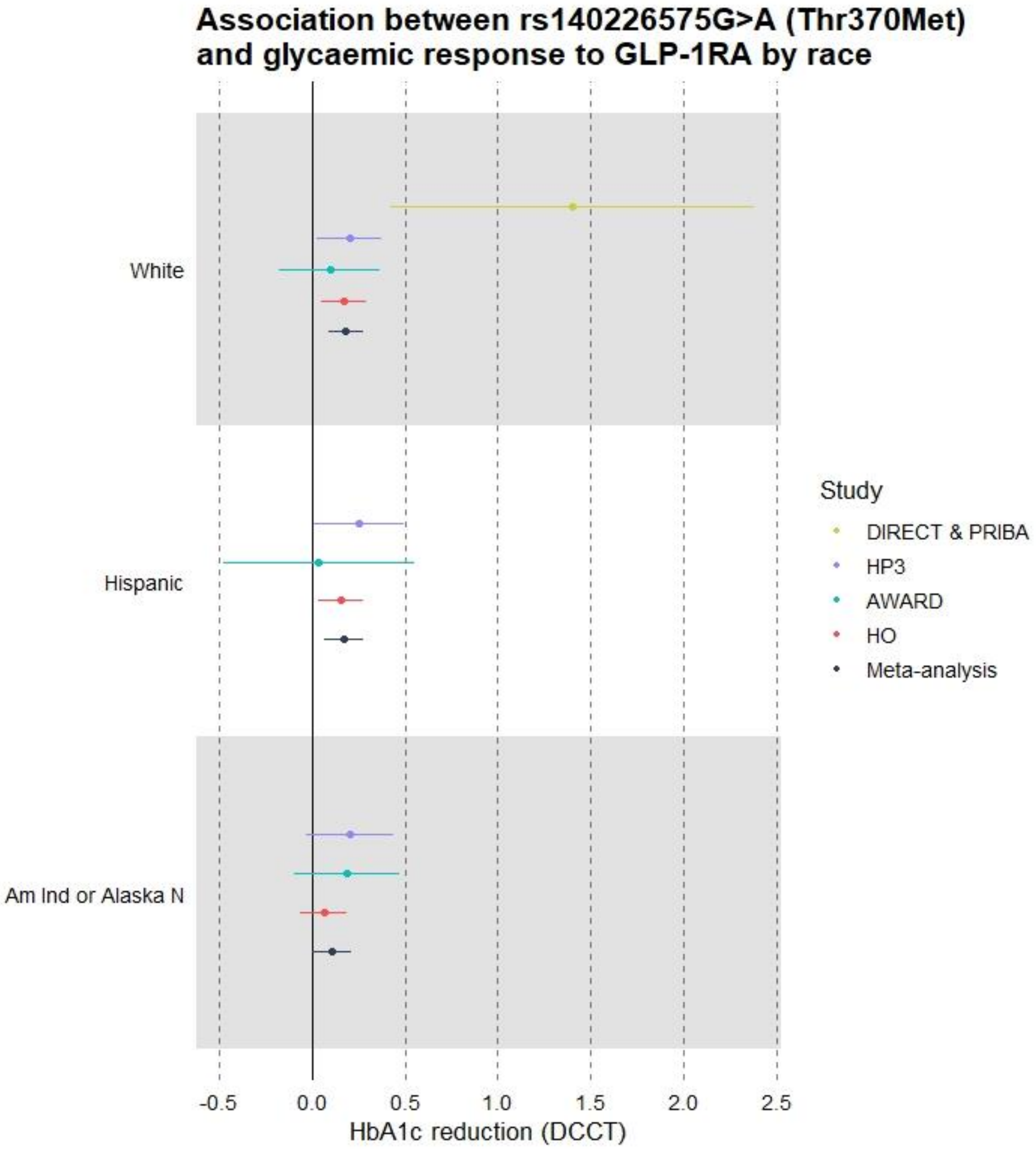


Supplementary Figure 4: Association between *ARRB1*-rs140226575G>A (Thr370Met) and HbA1c reduction after treatment with GLP-1RAs stratified by race. Effect estimates represent HbA1c reduction (DCCT) per minor allele. Adjustment was made for baseline HbA1c, age, sex, and first 3 principal components. DIRECT: The DIRECT (DIabetes REsearCh on patient stratification) study; PRIBA: The PRIBA (Predicting Response to Incretin Based Agents in Type 2 Diabetes) study; HP3: Harmony Phase 3 trials; AWARD: AWARD (Assessment of Weekly AdministRation of LY2189265 (dulaglutide) in Diabetes) trials; HO: Harmony Outcomes; Am Ind or Alaska N: American Indian or Alaska Native; DCCT: Diabetes Control and Complications Trial unit.
